# Interdependence of trabecular microarchitectural parameters from micro-computed tomography

**DOI:** 10.1101/2024.03.16.24304400

**Authors:** Nathan J. Neeteson, Annabel R. Bugbird, Lauren A. Burt, Steven K. Boyd

## Abstract

Standard microarchitectural analysis of bone using micro-computed tomography produces a large number of parameters that quantify the structure of the trabecular network. Analyses that perform statistical tests on many parameters are at elevated risk of making Type I errors. However, when multiple testing correction procedures are applied, the risk of Type II errors is elevated if the parameters being tested are strongly correlated. In this article, we argue that four commonly used trabecular microarchitectural parameters (thickness, separation, number, and bone volume fraction) are interdependent and describe only two independent properties of the trabecular network. We first derive theoretical relationships between the parameters based on their geometric definitions. Then, we analyze these relationships with an aggregated *in vivo* dataset with 2987 images from 1434 participants and a synthetically generated dataset with 144 images using principal component analysis (PCA) and linear regression analysis. With PCA, when trabecular thickness, separation, number, and bone volume fraction are combined, we find that 92% to 97% of the total variance in the data is explained by the first two principal components. With linear regressions, we find high coefficients of determination (0.827 – 0.994) and fitted coefficients within expected ranges. These findings suggest that to maximize statistical power in future studies, only two of trabecular thickness, separation, number and bone volume fraction should be used for statistical testing.

## 1 INTRODUCTION

### 1.1 Background

Micro-computed tomography (µCT) was established in the 1990s for the non-invasive measurement of 3D bone microarchitecture [1]. This eventually led to the development and proliferation of high-resolution peripheral quantitative computed tomography (HR-pQCT), which is an emerging medical imaging technology with a nominal spatial resolution of 60.7 µm that allows for direct, *in vivo*, quantitative analysis of cortical and trabecular microarchitecture [2]. HR-pQCT has been extensively applied to study how a wide variety of factors can affect the structure and strength of human bones at the smallest scales [3]–[5]. Increasingly, there is interest in the clinical utility of HR-pQCT, typically in the context of osteoporosis screening [6]. For example, several studies have shown that HR-pQCT images [7] and the derived strength and microarchitectural parameters [8]–[10] are significantly and independently predictive of fracture risk.

A typical HR-pQCT microarchitectural analysis includes the following quantitative trabecular parameters: bone mineral density (Tb.BMD), thickness (Tb.Th), separation (Tb.Sp), number (Tb.N), bone volume fraction (Tb.BV/TV), and inhomogeneity of trabecular network (Tb.1/N.SD). Sometimes it might also include structural model index (SMI), connectivity density (Conn.D), and degree of anisotropy (DA). Trabecular thickness, separation, number and bone volume fraction [11], [12] are based on the bone segmentation, as opposed to the direct gray-scale image, and are recommended as part of the minimum set to be reported [13]. Figure 1 depicts how these parameters are calculated and demonstrates how they are related geometrically.

**Figure 1:**
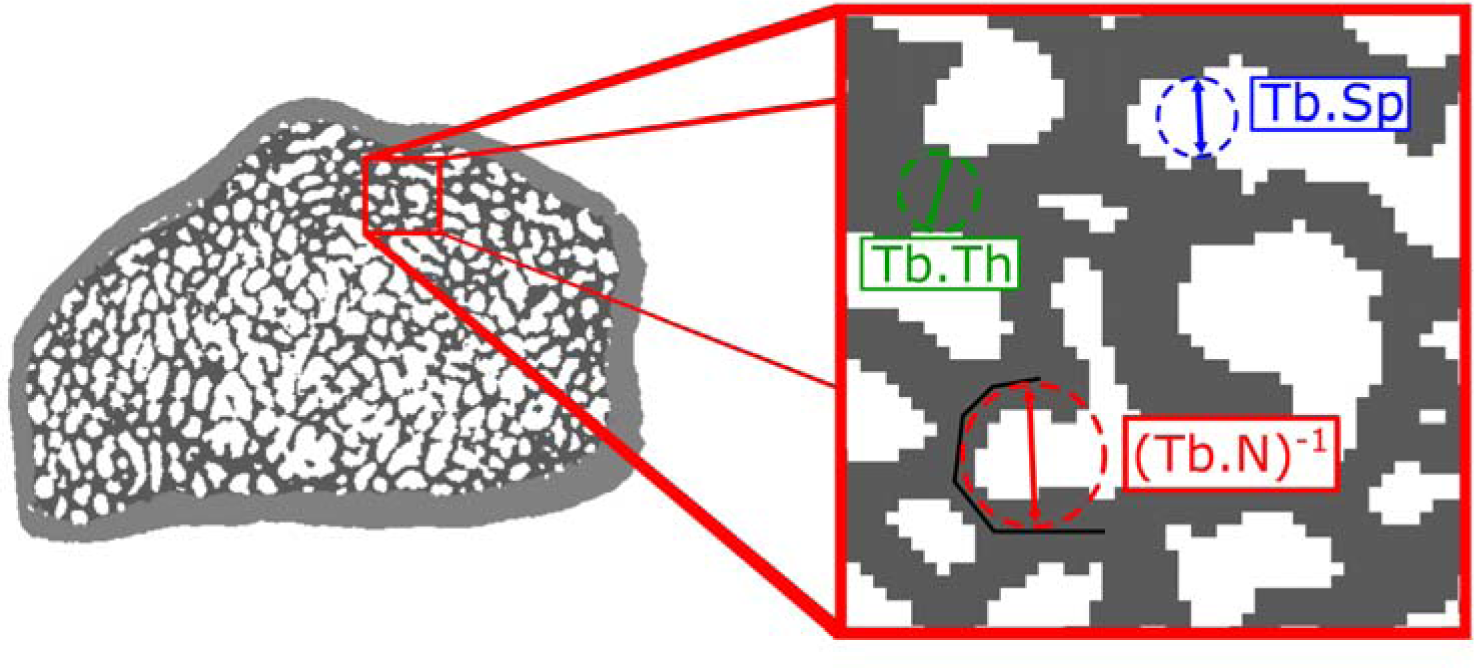
The trabecular microarchitectural parameters of focus are labelled on a sub-section of a distal radius HR-pQCT bone segmentation. Trabecular thickness, separation, and number are each calculated using Hildebrand’s thickness algorithm [11] on the bone segmentation, the background segmentation, and the background of the medial axis of the bone segmentation (black line), respectively. Bone volume fraction is the ratio of the number of voxels in the bone segmentation to the total number of voxels in the trabecular compartment [13].

These parameters are the basis for characterizing trabecular bone structure in cross-sectional studies, longitudinal studies, and by comparison to normative cohorts. However, a challenge of HR-pQCT is that there are many parameters available. To minimize both Type I and II errors, the selection of appropriate parameters is important. An insufficient set of parameters could result in physiologically relevant differences going undetected. Conversely, with many parameters, the problem of multiple testing arises, increasing the likelihood of Type I errors. Bonferroni correction [14] and Benjamini-Hochberg correction [15] are statistical methods that can be used to compensate for this risk, either by decreasing the effective significance thresholds proportionately to the number of hypotheses to be tested (Bonferroni) or by entirely modifying the statistical testing procedure (Benjamini-Hochberg). However, both procedures generally assume that the hypotheses being tested are independent. If there are redundant parameters, and therefore redundant hypotheses being tested, the tests will be more conservative than strictly necessary, increasing the risk of Type II errors and decreasing the statistical power of the analysis [16].

The objective of this study is to show that trabecular number and bone volume fraction are intrinsically linked to, and therefore statistically dependent on, thickness and separation. We demonstrate this dependence in both synthetic and *in vivo* microarchitectural data, using principal component analysis and linear regressions. These dependencies are important to recognize because they affect how to apply hypothesis testing and optimize study design.

### 1.2 Theoretical Relationships

We can observe in Figure 1 that the inverse of trabecular number is found by fitting spheres between the medial axis of the trabecular network, trabecular separation is found by fitting spheres between the trabecular surfaces, and trabecular thickness is found by fitting spheres within the trabecular surfaces [11]. Therefore, we can hypothesize that on average the difference between the inverse trabecular number and trabecular separation will be the trabecular thickness, or:

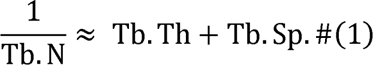

Turning to bone volume fraction, we first note that this parameter is defined as the ratio of bone volume to total volume within a compartment. We hypothesize that within a unit volume, this ratio will scale as some power of the ratio of the trabecular thickness to the sum of trabecular thickness and separation:

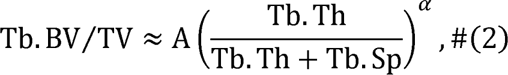

where *A* is a constant coefficient and *α* is the scaling power. If the trabeculae were axially aligned cylinders of uniform thickness and arranged in a grid with uniform spacing then *A* = 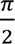 and *α* = 2 (Appendix A). If the trabeculae were evenly spaced, parallel plates with uniform thickness then *A*= 1 and *α* = 1. If the trabecular bone network includes both rod-like and plate-like geometry, as in typical *in vivo* data, we would expect 1 < *A* < 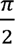 and 1 < *α* < 2. The natural logarithm transformation can be applied to each side of equation 2 and standard logarithm rules can be applied to produce a linearized relationship:

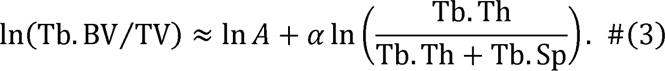

## 2 METHODS

### 2.1 Datasets

#### 2.1.1 *In vivo* Image Data

Image data captured *in vivo* with human participants were aggregated from four separate studies performed in our lab exploring bone microarchitecture by HR-pQCT, which were: a normative study [17], a hip fracture study [18], and two studies with high-performance athletes [19], [20]. All images were obtained using HR-pQCT (XtremeCT II, Scanco Medical AG, Brütisellen, Switzerland) with the standard fixed offset *in vivo* scan protocol at standard scan sites for the distal radius and tibia. In each image, 168 slices were collected with a nominal isotropic voxel size of 60.7 µm. All participants provided written informed consent prior to data collection, which was approved by the Conjoint Health Research Ethics Board at the University of Calgary. All images were analyzed following the standard semi-automated protocol for contouring and using the standard, manufacturer-implemented microarchitectural analysis.

#### 2.1.2 Synthetic Image Data

Synthetic bone segmentations were generated to explore the relationships between trabecular thickness, separation, number, and bone volume fraction when thickness and separation could be directly manipulated. The synthetic bone structures consisted of evenly spaced arrays of uniform cylinders aligned with the z-axis (Figure 2). Synthetic bone segmentations were generated with an isometric voxel size of 1 mm, an image size of 200 x 200 x 100 mm, trabecular radii ranging from 1 to 12 mm in increments of 1 mm, and unit grid spacing from 3 to 25 mm in increments of 1 mm. Constraining the grid spacing to be at least twice the trabecular radius plus one millimetre, so that synthetic trabeculae did not overlap, resulted in 144 total synthetic bone segmentations. Synthetic bone segmentations were generated and analyzed using manufacturer-provided software (Image Processing Language, IPL, version 5.42, Scanco Medical AG, Brütisellen, Switzerland).

**Figure 2:**
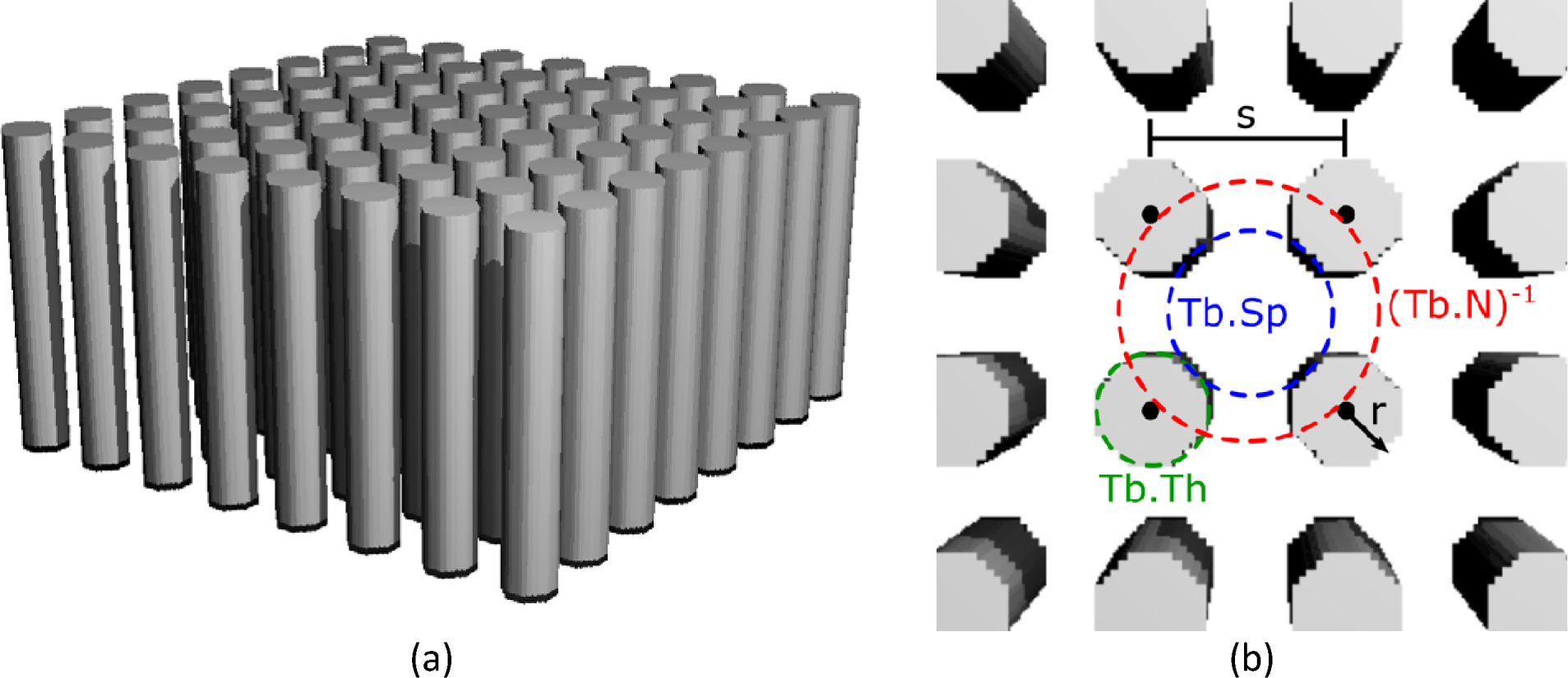
(a) Isometric rendering of a synthetic bone segmentation with a uniform cylinder radius of 7 mm and a uniform grid spacing of 25 mm. (b) Schematic diagram of the synthetic data labelling the grid spacing (s) and cylinder radius (r). Trabecular thickness, separation, and number measurements are approximately the diameter of the corresponding dashed circles.

### 2.2 Analysis

First, raw values for trabecular thickness, separation, number, and bone volume fraction were normalized within each data subset (synthetic, *in vivo*) by subtracting the mean and dividing by the standard deviation. For each dataset, principal component analysis (PCA) [21] was used to assess the interdependence of microarchitectural parameters by measuring the proportion of variance explained by each principal component, always using the normalized values. PCA was applied to trabecular thickness, separation, and number, then to thickness, separation, and bone volume fraction, and finally to all four parameters together. This process was repeated on each of the synthetic and *in vivo* data subsets. Appendix C describes and reports further analysis with additional semi-synthetic data generated from the *in vivo* data subset. PCA was also applied to trabecular thickness and separation together with the remaining parameters: structural model index, connectivity density, degree of anisotropy, and the inhomogeneity of the trabecular network.

Next, linear regressions were used to investigate how well equations 1 and 3 fit the synthetic and *in vivo* data. The coefficients of determination of these regressions were examined and the coefficients of the regressions with synthetic data were compared to the expected values to investigate the non-linear dependence of trabecular number and bone volume fraction on thickness and separation.

All analysis was performed using Python [22]. PCA models were created using scikit-learn [23] and linear regressions were created using statsmodels [24]. Visualizations were created using Inkscape [25], Matplotlib [26], Seaborn [27], and a manufacturer-provided 3D visualization tool (UCT3D, Version 4.2, Scanco Medical AG, Brütisellen, Switzerland).

## 3 RESULTS

Table 1 shows demographic information for all participants in the aggregated *in vivo* image dataset. For more information on the study inclusion and exclusion criteria and study-specific health data for these participants, please see the corresponding studies.

**Table 1:** Demographic information for the *in vivo* HR-pQCT data used in this work.

| Dataset | Number of participants | Median age [min, max] | Sex | Number of images |
| --- | --- | --- | --- | --- |
| Normative [17] | 1244 | 56 [18, 93] | 472M, 772F | 1243R, 1245T |
| Hip fracture [18] | 110 | 72 [57, 97] | 25M, 85F | 109R, 176T |
| Athletes [19], [20] | 80 | 24 [14, 41] | 36M, 44F | 103R, 111T |
| **Aggregated** | **1434** | **57 [14, 97]** | **533M, 901F** | **1455R, 1532T** |

Sex: M – male, F – female. Images: R – radius, T – tibia.

Figure 3 shows the results of PCA with trabecular thickness, separation, number, and bone volume fraction on the two data subsets. Similar trends are observed in both the *in vivo* and synthetic data subsets. Across each combination of parameters, the first two principal components combine to explain between 92 and 97% of the total variance.

**Figure 3:**
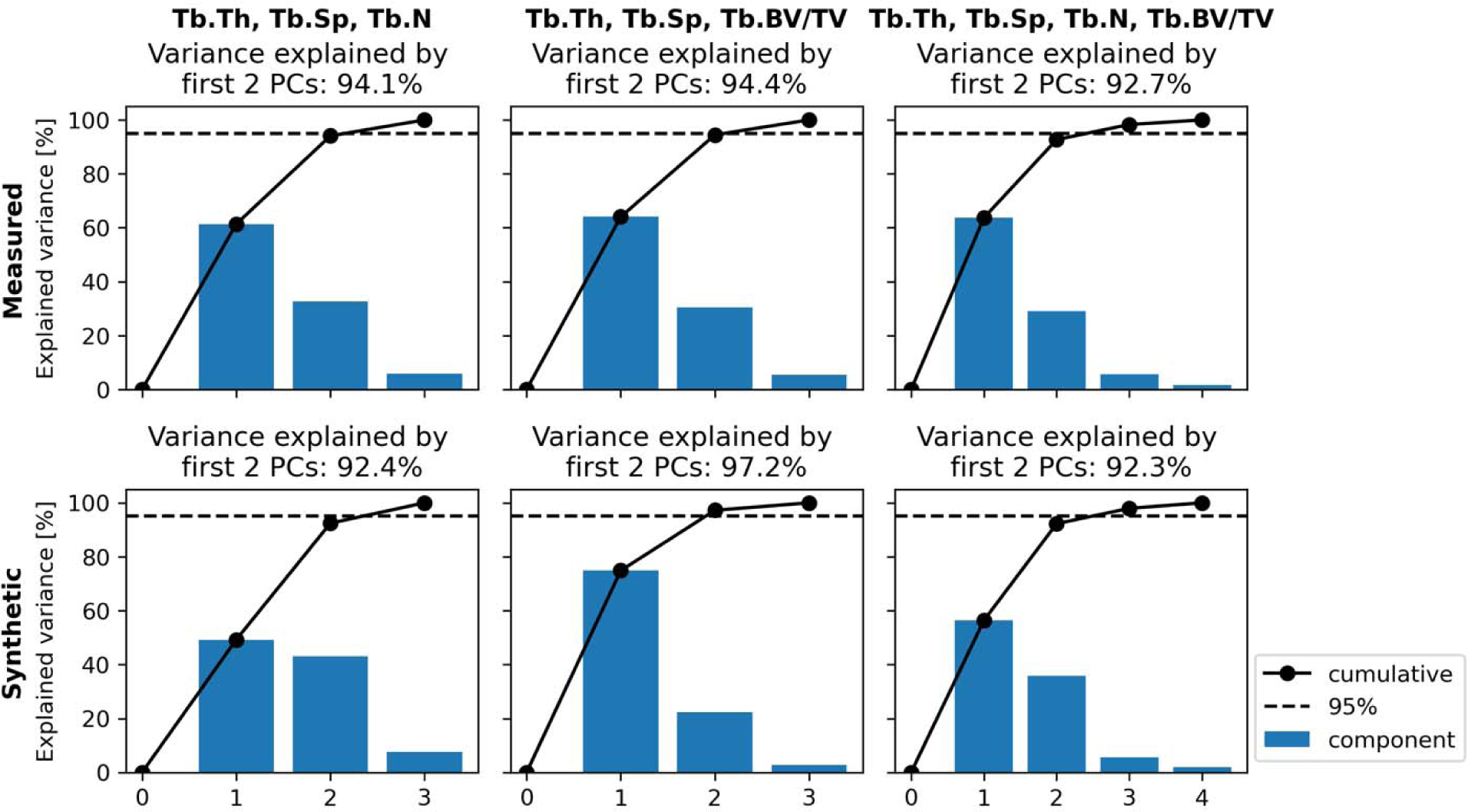
Explained variance for combinations of Tb.Th, Tb.Sp, Tb.N, and Tb.BV/TV. Measured: measurements in *in vivo* images from four aggregated studies. Synthetic: measurements in synthetically generated images.

Figure 4 shows the results of PCA with trabecular thickness, separation, structure model index, connectivity density, degree of anisotropy, and inhomogeneity of the trabecular network. For structure model index, connectivity density, and degree of anisotropy, the third principal component explains approximately 10% of the variance in the dataset. The inhomogeneity of the trabecular network is the exception: here, the first two principal components explain over 98% of the variance in the data. To assist with the qualitative interpretation of parameter interdependencies, pair plots are shown in Appendix B.

Table 2 shows the results of linear regressions of the form of equations 1 and 3 fit to the synthetic and *in vivo* datasets. All correlations were statistically significant, and all had coefficients of determination ≥0.98 except for Tb.BV/TV with the *in vivo* measured dataset, which was 0.827. The coefficients of trabecular thickness and separation for predicting the inverse of trabecular number were close to 1 with the synthetic data, as expected, while only the coefficient for trabecular separation was close to 1 with the *in vivo* data. In the regressions for the power-law scaling of Tb.BV/TV both intercepts were ≤0.45 and the scaling powers for the synthetic and *in vivo* data subsets were 1.81 and 1.27, respectively.

**Figure 4:**
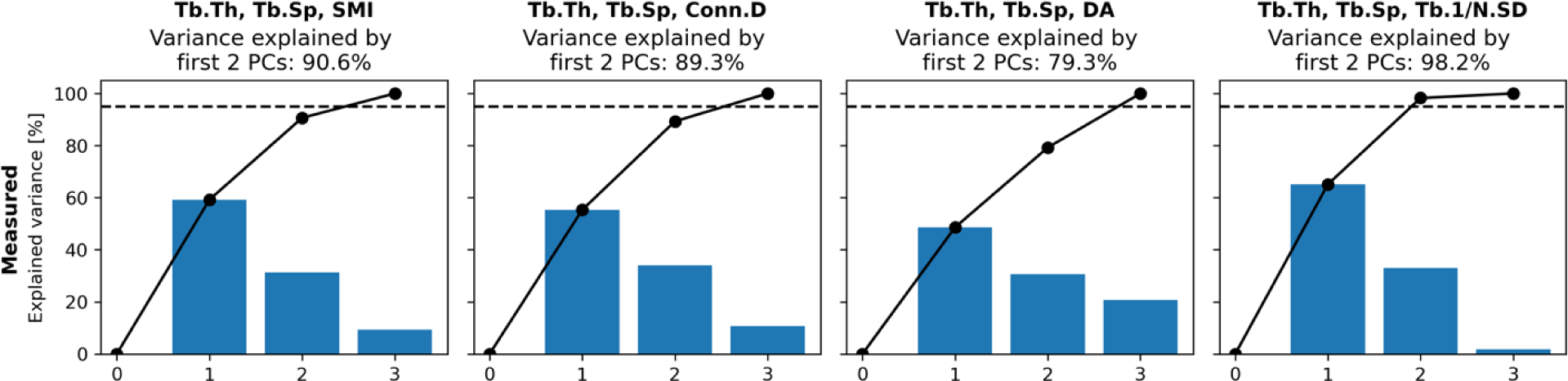
Explained variance for Tb.Th and Tb.Sp combined with SMI, Conn.D, DA, and Tb.1/N.SD individually, using the *in vivo* measured data subset.

**Table 2:**
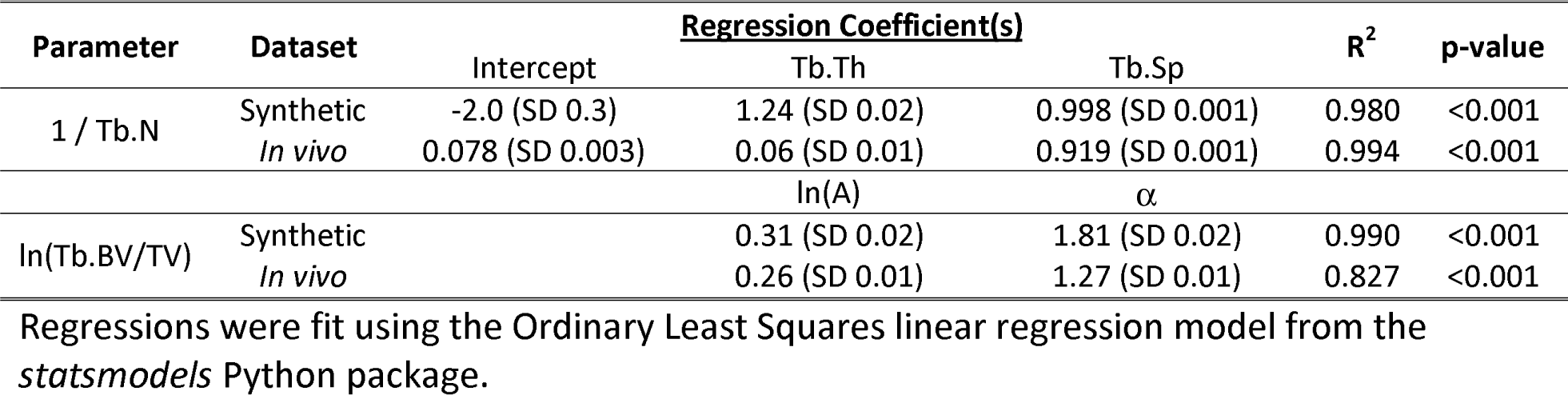
Predictor coefficients, coefficients of determination, and F-statistic-based p-values for linear regressions fit on relations of the form of equations 1 and 3 using the synthetic and measured datasets. Note that ln 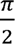 ≈ 0.45.

## 4 DISCUSSION

This study demonstrates through PCA, with both a large heterogeneous *in vivo* dataset and synthetic images, that when trabecular thickness, separation, number, and bone volume fraction are combined, 92% to 97% of the total variance is explained by only two underlying dimensions (Figure 3). This suggests that these parameters effectively describe only two orthogonal properties of the trabecular network. The implication is that only two of these parameters may be truly independent, while the other two are dependent, motivating further investigation into the specific hypothesized interdependencies.

When we fit linear regression models in the form of our theoretical approximate relationships to the synthetic and measured *in vivo* data (Table 2), we find that all correlations are statistically significant and have high coefficients of determination. We further find regression coefficients in line with theoretical expectations in the synthetic data. For the inverse of trabecular number, coefficients of trabecular thickness and trabecular separation are close to 1, as expected. For the bone volume fraction, the constant coefficient is approximately 1.37 (approximate expected value: 1.57) and the scaling power is 1.81 (expected value: 2). The regression coefficients deviated from expectations in the *in vivo* data. For the inverse of trabecular number, the coefficient of trabecular separation was close to 1, while the coefficient for trabecular thickness was 0.06. For the bone volume fraction, the constant coefficient was approximately 1.3 and the scaling power was approximately 1.3, in line with expectations for a mixture of rod-like and plate-like geometry. However, the regression for bone volume fraction in the *in vivo* data showed the weakest correlation, with a coefficient of determination of 0.827.

The deviation of the coefficients in the *in vivo* regression for inverse trabecular number data can be explained by the more complicated three-dimensional nature of the bone topology as compared to the synthetic data. The fact that the ratio of trabecular thickness to voxel size covered a much larger range in the synthetic data than in the *in vivo* data, 8 (SD 5) vs 4.1 (SD 0.5) (see Appendix B), could explain specifically why the dominant predictor for the inverse trabecular number in the in vivo dataset is trabecular separation, while the contribution of trabecular thickness seems to be negligible. However, we note that in all cases the regression coefficients are always significant and always positive, indicating that the trend of the dependent parameters will always be in the same direction given the same changes in the independent parameters.

In Figure 3, we show the proportion of explained variance when connectivity density, structural model index, the inhomogeneity of the trabecular network, and the degree of anisotropy are individually combined with trabecular thickness and separation. For all parameters except inhomogeneity of the trabecular network, we find that the proportion of variance explained by the third principal component is approximately 10%. In the case of inhomogeneity of the trabecular network, the proportion of variance explained by the third principal component is nearly negligible at <2%. However, we can contextualize this by examining the qualitative relationships between trabecular number, trabecular separation, and inhomogeneity of the trabecular network in the pair plot in Figure B2 (Appendix B). Here, it is apparent that the inhomogeneity of the trabecular network is proportional to both trabecular separation and the inverse trabecular number. The inverse trabecular number and inhomogeneity of the trabecular network are computed from the same local thickness field: as the mean and standard deviation, respectively [13]. Therefore, the dependence of the inhomogeneity of the trabecular network on trabecular thickness and separation can be described as an artifact of the dependence of the trabecular number on the trabecular thickness and separation.

Our findings have important implications for the use of microarchitectural parameters in clinical research, both when testing for the effects of an intervention on bone structure and when building predictive models. When using statistical tests to detect differences, it is important to carefully select the set of parameters to be tested. In the current guidelines, the minimum set of parameters to report includes trabecular thickness, separation, number, and bone volume fraction [13]. However, we have shown that trabecular number and bone volume fraction are fully dependent on trabecular thickness and separation. Accordingly, the change in any two of these parameters can be interpreted from the observed changes in the other two. For example: if trabecular separation increases significantly while trabecular thickness remains constant, it must be the case that bone volume fraction decreases and trabecular number decreases. Therefore, it is unnecessary to test the statistical significance of changes in more than two of these four parameters. The consequence of our findings is that one can increase the statistical power of their study design by selecting two of these four parameters to exclude from testing while losing no relevant information about geometric changes in the trabecular bone network.

However, we do not express a preference for which parameters should be kept or excluded from testing. Equations 1 and 2 can be manipulated to position any two parameters as the dependent variables, therefore study designers can select the two parameters of which the direct measurement will be the most relevant to their specific research question(s). These findings also do not preclude the dependent parameters from being tabulated and reported alongside the two selected for statistical testing, as they may be relevant for scientific interpretation. Finally, predictive models that incorporate trabecular microarchitectural parameters will likewise be adversely impacted by multicollinearity in the inputs [28]; therefore, we recommend that either only two of these four parameters be used or that dimensional reduction techniques, such as PCA, be used to reduce these four parameters to two dimensions as part of data preprocessing.

In this work, we have focused our analysis on trabecular thickness, separation, number, and bone volume fraction for linear regression analysis, while reporting only the explained variance in PCA for connectivity density, structural model index, the inhomogeneity of the trabecular network, and the degree of anisotropy combined with trabecular thickness and separation. There are two motivations for this: First, we have focused our attention on those trabecular parameters that are both based on the bone segmentation and recommended as the minimum set to be reported in the most recent guidelines for HR-pQCT analysis in adults [13]. Second, we have focused on those parameters that are based on either a simple ratio of volumes (bone volume fraction) or the mean of a local thickness field calculated via Hildebrand’s algorithm [11] (thickness, separation, number). The definitions of these parameters are simple to understand by looking at the diameters of circles superimposed on cross-sections of trabeculae, such as Figures 1 and 2(b). Correspondingly, it is relatively simple to derive the expected relationships of trabecular number and bone volume fraction on trabecular thickness and separation using only geometric arguments. This same simple geometric analysis does not seem to work for any of connectivity density, structural model index, the inhomogeneity of the trabecular network, or the degree of anisotropy.

There are important limitations to the analysis that should be noted. First, while our *in vivo* data contains a large sample of images and individuals (3152 total images from 559 men and 931 women) from a clinically diverse set of studies, the geographic and ethnic diversity of our data is limited: all study data was collected in Calgary, Alberta, Canada and the overwhelming majority of study participants were White. However, our hypotheses and arguments are based primarily on the geometric definitions of the parameters and how they relate to each other, and we believe our findings are fundamental to the measurement methodology. Therefore, we have no cause to hypothesize that our results would fail to replicate in a separate cohort with more geographic and ethnic diversity. Second, PCA is intended for the discovery of linear relationships among variables in a dataset, while here we have used it to investigate potential non-linear relationships. However, we have used PCA in this context only for exploration and to motivate the investigation of highly specific non-linear interdependencies via linear regression analysis on linearized forms of those relationships.

We have demonstrated that only two of trabecular thickness, separation, number, and bone volume fraction are independent parameters of the trabecular network. When performing statistical testing, only two of these parameters should be used, to minimize type I or II errors (depending on whether multiple-testing correction is applied, as is recommended). Likewise, only two of these parameters should be included in the input space when designing predictive models that incorporate microarchitectural parameters.

## Data Availability

The raw image data used in this study are not publicly available.

## ACKNOWLEDGEMENTS

The authors acknowledge the support and contributions of the staff and students of the Bone Imaging Laboratory at the University of Calgary, Dr. Danielle Whittier for curating the normative study data, and the recruitment efforts of the Calgary Fracture Liaison Service Nurses, the Orthopaedic Trauma Research, and the Gastroenterology Clinic/Endoscopy Unit team at Foothills Medical Centre in Calgary at Foothills Medical Centre, as well as the collaborative efforts of the Canadian Sport Institute Calgary. This work was supported financially by the Canadian Institutes of Health Research (CIHR) [PJT 162189] and The Arthritis Society (TAS) PhD Salary Award [TGP-21-0000000093].

## CODE AVAILABILITY

All code used for generating synthetic data and analysis of all datasets is available in the following GitHub repository: https://github.com/Bonelab/uct-parameter-interdependence

## DATA AVAILABILITY

The raw images used in this study are not available, to protect the privacy and confidentiality of the participants.

## APPENDIX

## Appendix A

### – Expected relationship between Tb.BV/TV, Tb.Th, and Tb.Sp in synthetic data

**Figure A1:**
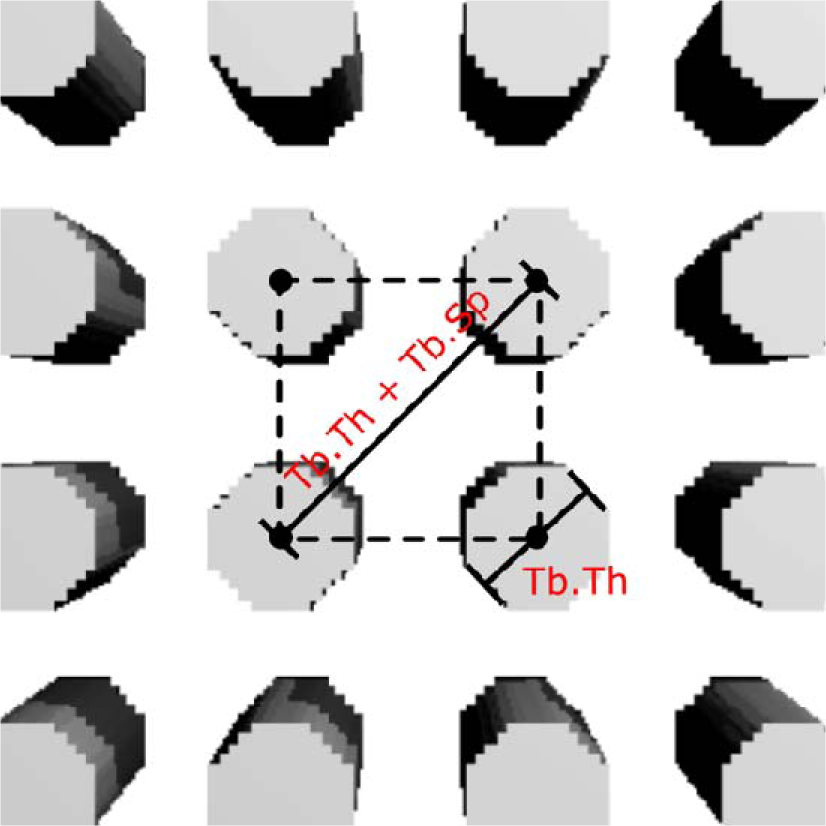
Axial schematic of the synthetic data showing the repeating unit cube that composes the entire volume. The diagonal of the cube’s axial face and the cylinder diameters are labelled in terms of microarchitectural parameters.

Figure A1 shows a schematic of the repeating unit cube that composes the entire volume of any given synthetic bone segmentation used in this work. The diagonal of this cube is the trabecular thickness and separation combined. Since we are eventually interested in the ratio of two volumes, we may assume the cube has unit depth without loss of generality. Therefore, the total volume of this cube, as a function of trabecular thickness and separation, is:

Each unit cube contains one quarter of a trabecula at the corners. Since all trabeculae are identical and the diameter of a trabecula is the trabecular thickness, by the definition of the volume of a spherical cylinder and again assuming unit depth, the total volume of bone within the unit cube is:

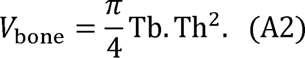

Combining equations (A1) and (A2) with the definition of bone volume fraction generates a theoretical expectation for the bone volume fraction in the synthetic data:

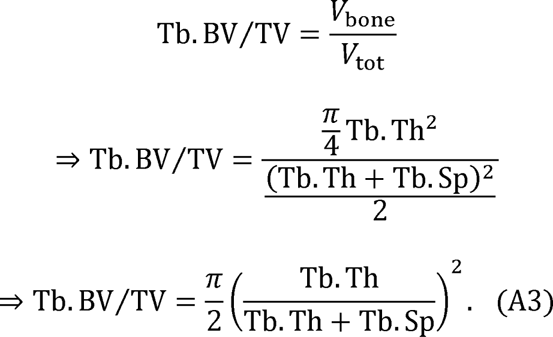

This relationship should hold exactly in the synthetic data, and a similar trend should be observed in *in vivo* data.

## Appendix B

### – Pair plots

**Figure B1:**
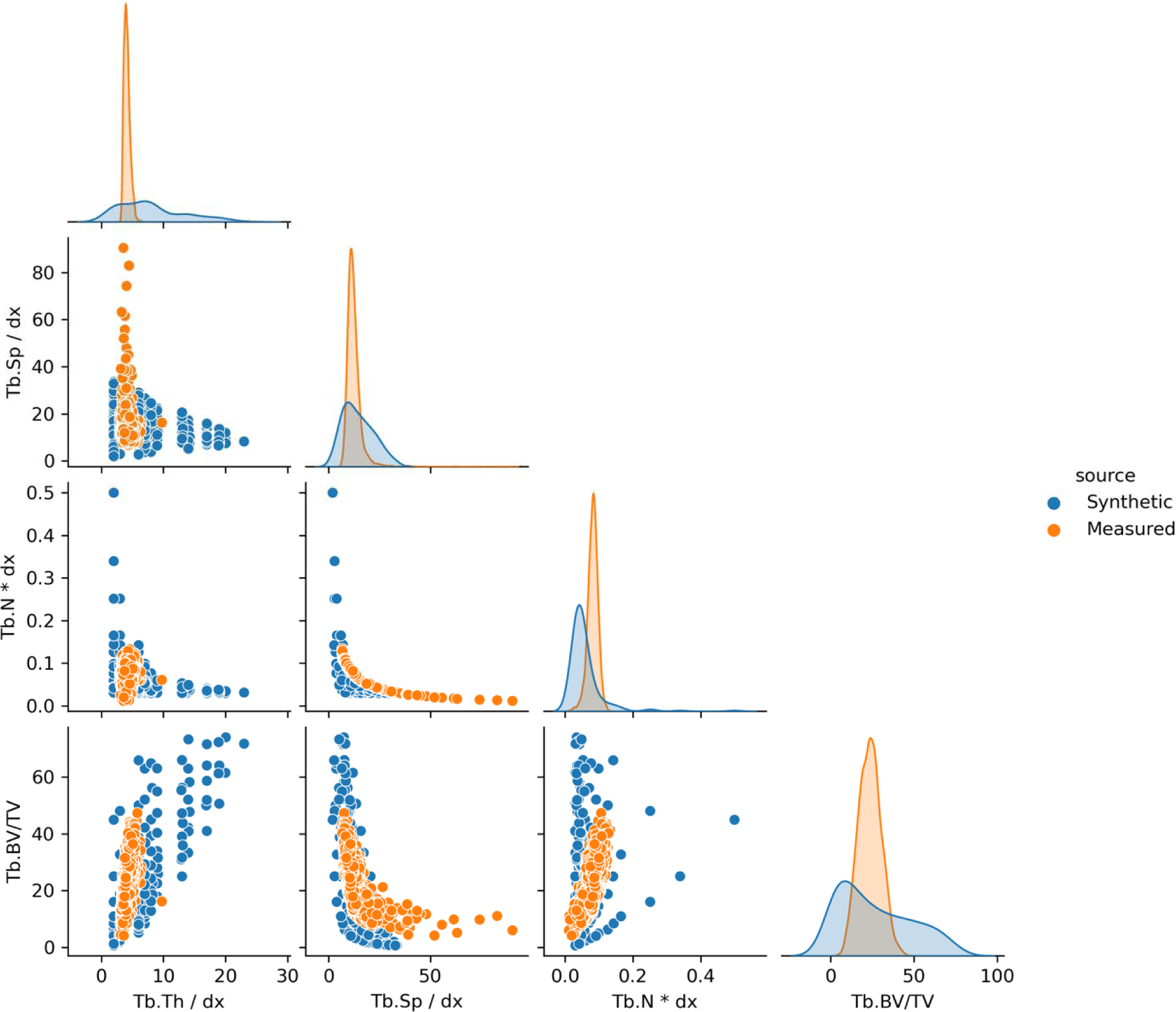
Pair plot for normalized trabecular thickness, separation, number, and bone volume fraction for the synthetic and *in vivo* data. Trabecular thickness and separation are normalized by dividing by voxel size, and trabecular number is normalized by multiplying by voxel size. Normalization is performed to allow comparison between datasets. Off-diagonal plots show bivariate scatterplots, while diagonal plots show univariate kernel density estimate plots. The means and standard deviations of the normalized trabecular thicknesses are 8 (SD 5) and 4.1 (SD 0.5) voxels in the synthetic and *in vivo* (Measured) datasets, respectively.

**Figure B2:**
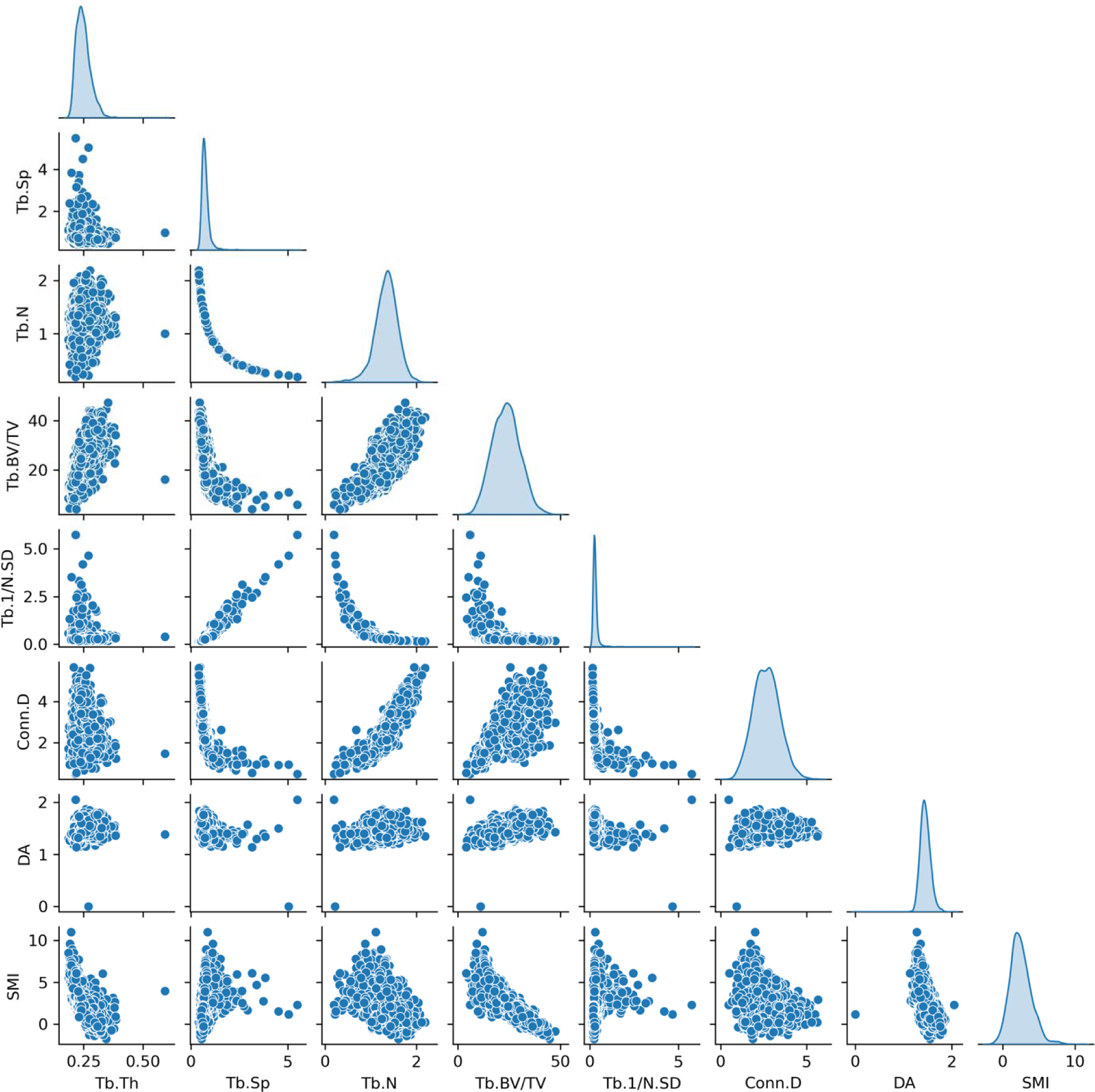
Pair plot for all trabecular microarchitectural parameters, with *in vivo* data only.

## Appendix C

### – Explained variance with PCA with additional semi-synthetic datasets

Further demonstrating the theoretical interdependence described by equations 1 and 2, two additional semi-synthetic datasets were constructed. First, a “dependent” dataset was constructed using trabecular thickness and separation from the measured *in vivo* data as inputs to equations 1 and 2 (with *A* = 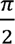 *α* = 2) and generating synthetic dependent values for trabecular number and bone volume fraction. Second, an “independent” dataset used the measured *in vivo* trabecular thickness and separation but fully randomly generated synthetic trabecular number and bone volume fraction values. These were generated by randomly sampling values from normal distributions with means and standard deviations equal to those observed in the *in vivo* distributions of trabecular number and bone volume fraction.

**Figure C1:**
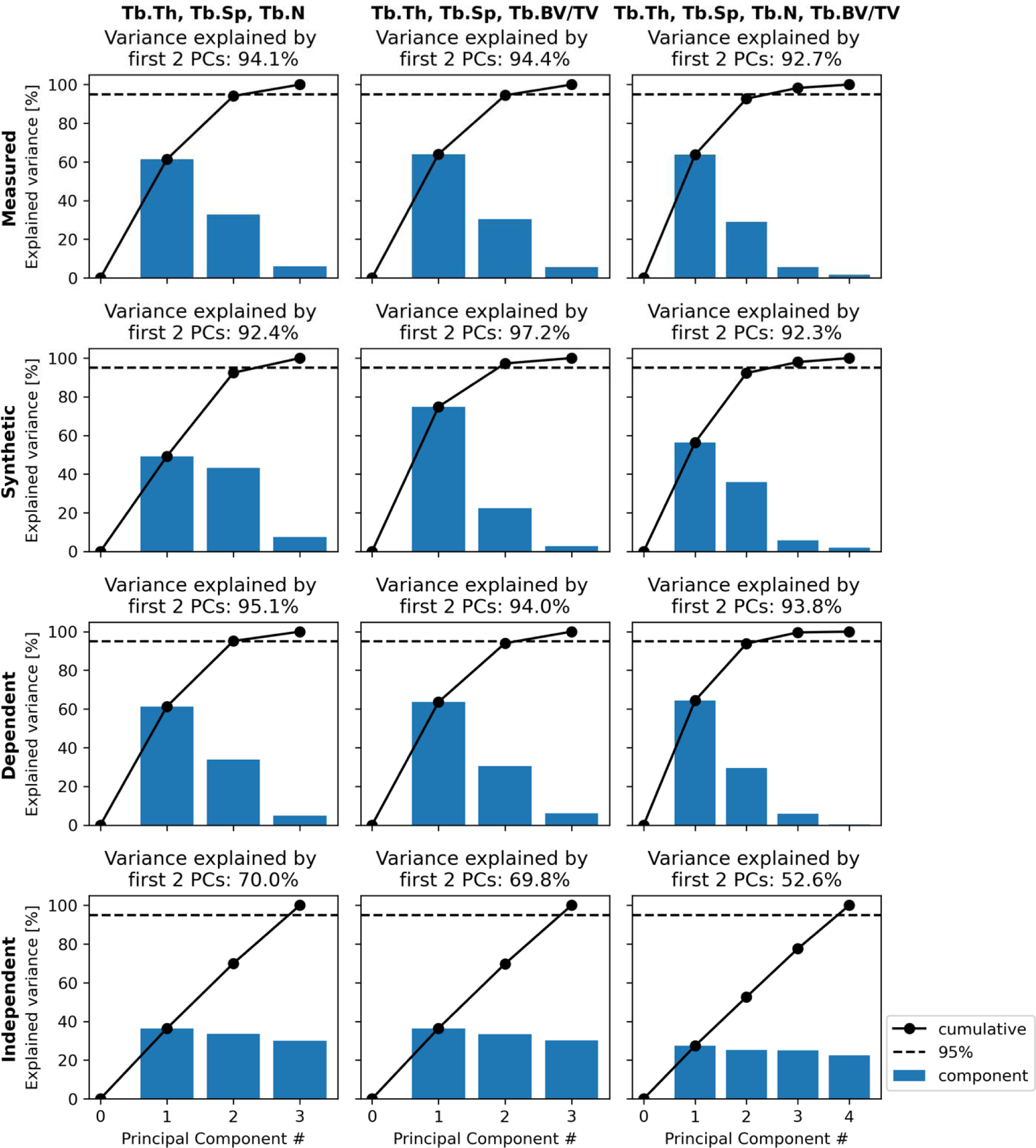
Explained variance for combinations of Tb.Th, Tb.Sp, Tb.N, and Tb.BV/TV for four distinct datasets. Synthetic: measurements in synthetically generated images. Measured: measurements in *in vivo* images from human participants across five studies. Dependent: Using the measured *in vivo* Tb.Th and Tb.Sp, Tb.N and Tb.BV/TV are generated using the hypothesized proportionalities in equations 1 and 2. Independent: Tb.Th and Tb.Sp are from *in vivo* measurements, while Tb.N and Tb.BV/TV are randomly generated to ensure statistical independence from Tb.Th and Tb.Sp.

## Notes

### Competing Interest Statement

The authors have declared no competing interest.

### Author Declarations

The Conjoint Health Research Ethics Board of The University of Calgary gave ethical approval for this work.

